# Visual and pharmacotherapy outcomes after transverse sinus stenting for idiopathic intracranial hypertension

**DOI:** 10.1101/2023.10.23.23297230

**Authors:** Armin Handzic, Jim Shenchu Xie, Eef Hendriks, Pascal Mosimann, Patrick Nicholson, Jonathan Micieli, Edward Margolin

**Affiliations:** University of Toronto, Faculty of Medicine, Department of Ophthalmology and Vision Sciences, Toronto, Ontario, Canada; Michael G. DeGroote School of Medicine, McMaster University, Hamilton, Ontario, Canada; University of Toronto, Faculty of Medicine, Department of Medical Imaging, Division of Neuroradiology, Toronto, Ontario; University of Toronto, Faculty of Medicine, Department of Medicine, Division of Neurology, Toronto, Ontario, Canada; Kensington Vision and Research Center, Toronto, Ontario, Canada

**Keywords:** idiopathic intracranial hypertension, stenting, venous sinus

## Abstract

**Background:** Transverse sinus stenting (TSS) is an increasingly commonly used treatment for patients with idiopathic intracranial hypertension (IIH). However, detailed neuro-ophthalmic evidence on visual and pharmacotherapy outcomes after TSS is scarce and heterogeneous. This study aimed to describe the visual outcomes of patients undergoing TSS for IIH and to ascertain the proportion of patients who could be weaned off intracranial pressure (ICP)-lowering medication after this procedure.

**Methods:** A retrospective chart review of all patients with IIH from two tertiary academic neuro-ophthalmology practices who underwent TSS between 2016 and 2022 was performed. Indications for stenting included failure of pharmacotherapy, intolerance of pharmacotherapy, and acute vision loss from severe papilledema. Data on demographics, symptoms, visual function, and TSS were collected. The paired Wilcoxon rank sum test was used to compare changes in visual acuity (VA) and visual field mean deviation (VFMD) between the baseline and most recent visits.

**Results:** Of 435 patients with IIH, 15 (13 women) met inclusion criteria. After TSS, ICP-lowering pharmacotherapy was discontinued in 10 patients and decreased in 4; one patient was not on ICP-lowering medication before TSS. All patients experienced resolution or improvement of symptoms (10 resolution, 4 improved, 1 asymptomatic before TSS) and papilledema (11 resolution, 4 improved) after stenting. Papilledema resolution was confirmed with optical coherence tomography-measured peripapillary nerve fibre layer thickness (median decrease 147 μm, interquartile range 41.8 – 242.8, p<0.001). Change in VA between the baseline and most recent visit was not significant, but VFMD improved significantly after stenting (median increase 3.0, IQR 2.0 – 4.2, p<0.001). No patient developed transverse sinus restenosis nor in-stent thrombosis postoperatively across a median venogram follow-up of 20.8 weeks (11.3 – 49.8) and no patient required subsequent surgical intervention for IIH.

**Conclusion:** In this cohort of patients with IIH and fulminant presentation, medication resistance, or medication intolerance, TSS was an effective and safe treatment modality. Most patients were able to stop ICP-lowering medications while demonstrating striking improvement in symptomatology and visual function.

## INTRODUCTION

Stenting of transverse sinus stenosis is becoming an increasingly frequently used surgical treatment for idiopathic intracranial hypertension (IIH). While the exact pathogenic mechanism of IIH remains unclear, it has become evident that patients with IIH develop stenosis along the transverse sigmoid sinus junction, either bilaterally or in one dominant sinus.^1^ This finding can be assessed with 3D contrast-enhanced magnetic resonance imaging (MRI) and is highly sensitive and specific for IIH.^2^

First-line treatment for IIH consists of weight loss and acetazolamide, which has been validated in a large multicenter trial of patients with mild visual field (VF) loss.^3,4^ Notably, up to 2.9% of patients with IIH present with fulminant vision loss and up to 25% develop permanent visual impairment due to papilledema.^5,6^ Furthermore, approximately 10% of patients are refractory to conservative treatment and require surgical intervention to reduce intracranial pressure (ICP).^7^ In addition to transverse sinus stenting (TSS), other commonly performed procedures for IIH include cerebrospinal fluid (CSF) shunt placement and optic nerve sheath fenestration (ONSF).

While several systematic reviews and meta-analyses have investigated post-TSS visual acuity, papilledema, symptomatology, complications, and IIH recurrence, none have examined visual field, optical coherence tomography (OCT), and pharmacotherapy outcomes.^8–12^ This study aimed to determine the proportion of patients treated with TSS who could be weaned off ICP-lowering pharmacotherapy and to describe their detailed neuro-ophthalmic outcomes.

## Methods

### Study design, setting and participants

We performed a retrospective chart review of all patients at two neuro-ophthalmology tertiary centres between 2016 and 2022 who underwent TSS for IIH. IIH diagnosis was made by two fellowship-trained neuro-ophthalmologists (EM, JM) according to the modified Dandy criteria.^1^ We excluded patients with an identifiable cause for increased ICP, a condition other than IIH that caused vision loss, or less than 1.5 months of follow-up after stenting. Unreliable visual field (VF) tests, defined as false positives >25%, false negatives >30%, or fixation losses >25%, were excluded from data analysis.

Indications for TSS included failure of pharmacotherapy, intolerance of ICP-lowering medications, and fulminant IIH at presentation. Failure of pharmacotherapy was defined as persistence of papilledema and/or continuous visual deterioration despite a maximal daily dose of acetazolamide 3 g or topiramate 200 mg. Fulminant IIH presentation was defined as initial best-corrected visual acuity (VA) worse than 20/40 and/or an initial VF mean deviation (VFMD) worse than -5 dB. This study was approved by the University of Toronto Health Sciences Research Ethics Board and adhered to the tenets of the Declaration of Helsinki.

### Study procedure and data collection

All patients underwent comprehensive neuro-ophthalmologic evaluation by a fellowship-trained neuro-ophthalmologist (EM, JM) that included a thorough history, Snellen VA measurement, biomicroscopic examination, VF testing, and peripapillary OCT. Humphrey VF testing (24-2 algorithm) was performed using the Swedish Interactive Thresholding Algorithm (SITA) Fast protocol. A Cirrus spectral-domain OCT unit (Zeiss, Dublin, California, USA) was used to measure the thickness of the peripapillary retinal nerve fibre layer (pRNFL). MRI and magnetic resonance venography (MRV) of the brain were carried out in all patients before TSS. We obtained clinical data from the baseline and most recent follow-up visits as well as interventional data from the stenting report.

### Treatment specifications

TSS was performed under general anesthesia in supine position using a biplane neuroangiography system (Philips Allura Xper FD20/20, Eindhoven, The Netherlands). Patients were preloaded with aspirin 81 mg and clopidogrel 75 mg daily for 5 days prior to TSS. First, a cerebral angiography was obtained via 5 French (Fr) radial artery access to visualize the arterial and venous anatomy of the cerebral and cerebellar hemispheres, including a venous 3D rotational angiography of the dural venous sinus. Second, via an 8 Fr common femoral or jugular vein access, pressure measurements were performed with a microcatheter (Excelsior SL-10, Stryker Neurovascular, Hamilton, Ontario, Canada) in the dural venous system proximal and distal to the stenotic segment. Third, after ensuring a trans-stenotic pressure gradient of greater than 5 cm of H_2_O, a carotid wall stent measuring 7 × 40 mm (Boston Scientific, Mississauga, Ontario, Canada) was deployed across the stenotic segment, followed by repeat pressure measurements. A 3-month follow-up CTV brain was routinely performed, and double antiplatelet therapy was continued for at least 3 months.

### Outcomes

The primary outcome was the proportion of patients for whom ICP-lowering medications were discontinued following TSS. Secondary outcomes included changes in symptomatology, visual function, papilledema severity, and pRNFL thickness between the baseline and most recent follow-up visits. We also examined rates of stenting failure, transverse sinus restenosis, and postoperative complications. Stenting failure was defined as the persistence of papilledema or medication use at the final follow-up. Symptoms considered to indicate raised ICP included headache, pulsatile tinnitus, diplopia secondary to abducens nerve palsy, and transient visual obscurations.

### Data analysis

Data were summarized using frequencies and percentages for categorical variables and medians and interquartile ranges (IQR) for continuous variables. The paired Wilcoxon rank sum test with continuity correction was used to compare changes in VA, VFMD, and pRNFL thickness between the baseline visit and most recent visit. The unpaired Wilcoxon rank sum test was used to evaluate whether duration of IIH and duration of symptoms before TSS differed between patients with and without stenting failure. Additionally, the Kendall rank correlation coefficient was used to determine whether IIH and visual symptom duration were associated with visual recovery (i.e. change in VA and change in VFMD). We considered a VA change of ≥0.1 logMAR and a VFMD change of ≥3 dB to be clinically significant. Data analysis was performed using R (version 4.2.1; R Foundation for Statistical Computing, Vienna, Austria). All statistical tests were two-sided and *P* values <0.05 were considered statistically significant.

## Results

Of 435 patients with IIH, 30 eyes of 15 patients (13 female) met inclusion criteria. The demographic, clinical, and interventional characteristics of included patients are summarized in Table 1. Patients had a median age of 27 years (IQR: 22 – 32), body mass index of 35.2 kg/m^2^ (30.8 – 41.1), and opening pressure of 42 mm of H_2_O (36 – 49). Three patients (20%) underwent surgical intervention to lower ICP before TSS: patient 1 underwent left ONSF and had recurrence of papilledema; patient 4 underwent two lumboperitoneal shunt placements during their childhood, and since papilledema recurred in their 30s, we assumed interval development of shunt dysfunction; and patient six underwent lumboperitoneal shunt placement which was displaced at presentation. All but one patient experienced ICP-related symptoms before stenting. Indications for surgical intervention were pharmacotherapy resistance in 9 patients, medication intolerance in 2 patients, and fulminant presentation in 4 patients. The median time between IIH diagnosis and TSS was 26.4 weeks (3.9 – 143.8), and the median duration of ICP-related symptoms before TSS was 6.7 weeks (3.2 – 12.2). TSS was unilateral in 13 patients. Stent insertion produced a median trans-stenotic gradient reduction of 12 mm Hg (9.5 – 20.0).

**Table 1.** Patient demographics and treatment characteristics.

| Patient | Age (y)/<br>Sex | Comorbidities | Previous IIH-related<br>surgeries | BMI<br>(kg/m<sup>2</sup>) | LP OP (cm<br>of H<sub>2</sub>O) | Indication for<br>stent | Time from IIH<br>diagnosis to<br>stent (weeks) | Stent<br>laterality | Stenosis grade | Stent<br>location |
| --- | --- | --- | --- | --- | --- | --- | --- | --- | --- | --- |
| 1 | 70s/M | HTN, DLD | Left ONSF | 39.4 | 42 | Refractory to<br>medications | 78.1 | Left | Severe | TS-SS |
| 2 | 20s/F | Celiac disease, MDD, BPD,<br>mastectomy | None | 30.8 | 42 | Refractory to<br>medications | 209.4 | Right | Moderate | TS |
| 3 | 30s/F | None | None | 41.1 | 44 | Fulminant<br>presentation | 1 | Right | Moderate | TS |
| 4 | 40s/M | Congenital hydrocephalus | 2 LP shunts | 34.7 | 36 | Refractory to<br>medications | 26.4 | Left | Severe | TS |
| 5 | 40s/F | None | None | 29.4 | 28 | Intolerant of<br>medications | 76.1 | Right | Severe | TS-SS |
| 6 | 20s/F | None | LP shunt but<br>displaced | 38 | 50 | Refractory to<br>medications | 275.6 | Bilateral | Moderate right,<br>severe left | TS-SS<br>bilaterally |
| 7 | 30s/F | None | None | 52.2 | 36 | Refractory to<br>medications | 279.3 | Left | Severe | TS-SS |
| 8 | 20s/F | MDD | None | 45.6 | 58 | Fulminant<br>presentation | 1.7 | Bilateral | Severe right,<br>moderate left | TS right, TS-SS<br>left |
| 9 | 20s/F | Iron-deficiency anemia, pre-<br>eclampsia | None | 42.5 | 24 | Refractory to<br>medications | 77.1 | Right | Severe | TS-SS |
| 10 | Teens/F | None | None | 35.2 | 49 | Fulminant<br>presentation | 2.6 | Right | Severe | TS |
| 11 | 20s/F | None | None | 29.2 | 36 | Fulminant<br>presentation | 2.1 | Right | Severe | TS-SS |
| 12 | 20s/F | Breast fibroadenoma | None | NR | 48 | Refractory to<br>medications | 5.1 | Right | Mild | TS-SS |
| 13 | 30s/F | Nephrolithiasis | None | 22.6 | 23 | Intolerant of<br>medications | 14.7 | Left | Severe | TS |
| 14 | 20s/F | PCOS, OSA | None | 34.6 | 50 | Refractory to<br>medications | 337 | Right | Mild | TS |
| 15 | 20s/F | Ehlers Danlos syndrome treated<br>with suboccipital and cervical<br>spine decompression | None | NR | 36 | Refractory to<br>medications | 11 | Right | Severe | TS-SS |

BMI = body mass index, BPD = bipolar disorder, DLD = dyslipidemia, HTN = hypertension, IIH = idiopathic intracranial hypertension, LP = lumboperitoneal, MDD = major depressive disorder, NR = not recorded, OP = opening pressure, OSA = obstructive sleep apnea, PCOS = polycystic ovarian syndrome, SS = sigmoid sinus, TS = transverse sinus

Patients were followed for a median of 66.9 weeks (36.1 – 109.3). Table 2 summarizes each patient’s course of ICP-related symptoms, papilledema, and pharmacotherapeutic management. After TSS, ICP-lowering medications were discontinued in 10 patients (71%) and decreased in 4 patients (29%). Patient 10 presented to our neuro-ophthalmology centre after TSS and was not prescribed ICP-lowering medication preoperatively. All 4 patients on decreased doses of ICP-lowering medication were in the process of weaning off acetazolamide; patient 2 remained on 1 g of acetazolamide daily for psychological reassurance rather than clinical need. All patients experienced postoperative resolution or improvement of symptoms (10 [71%] resolution, 4 [29%] improved) and papilledema (11 [73%] resolution, 4 [27%] improved). Specifically, headache resolved in 10 of 13 patients (77%), pulsatile tinnitus in 8 of 10 patients (80%), diplopia in 4 of 6 patients (67%), and transient visual obscurations in 2 of 4 patients (50%).

**Table 2.** Symptomatology, papilledema, and pharmacotherapy before and after stenting.

| Patient | Total FU (weeks) | Symptoms |  | Papilledema – right eye |  | Papilledema – left eye |  | Medications (daily dose) |  |
| --- | --- | --- | --- | --- | --- | --- | --- | --- | --- |
|  |  | Pre | Post | Pre | Post | Pre | Post | Pre | Post |
| 1 | 73 | None | None | Moderate | Resolved | Mild | Resolved | ACZ 1 g | Stopped |
| 2 | 49.1 | HA, PT | HA resolved, PT decreased | Mild | Resolved | Moderate | Resolved | ACZ 2.5 g, TPM 200 mg | ↓ to ACZ 1 g |
| 3 | 65.9 | HA, PT | Completely resolved | Severe | Resolved | Severe | Resolved | ACZ 3 g | Stopped |
| 4 | 96.4 | PT | Completely resolved | Severe | Resolved | Severe | Resolved | ACZ 2 g | Stopped |
| 5 | 55.3 | PT | Completely resolved | Severe | Mild | Severe | Mild | None | Stopped |
| 6 | 220.1 | HA, PT | Completely resolved | None | Resolved | None | Resolved | ACZ 4 g | Stopped |
| 7 | 108.4 | HA, PT, DP | Completely resolved | Severe | Mild | Severe | Mild | ACZ 2 g | Stopped |
| 8 | 110.1 | HA, PT | PT resolved, HA decreased | Severe | Resolved | Severe | Resolved | ADZ 4 g | ↓ to ACZ 500 mg |
| 9 | 174.6 | HA, PT, TVO, DP | All decreased | Severe | Mild | Severe | Mild | ACZ 3 g | ↓ to ACZ 500 mg |
| 10 | 7.4 | HA, TVO, DP | Completely resolved | Severe | Mild | Severe | Mild | None | None |
| 11 | 23.0 | HA, DP | Completely resolved | Severe | Resolved | Severe | Resolved | ACZ 2 g | ↓ to ACZ 750 mg |
| 12 | 17.1 | HA, PT, DP | Completely resolved | Severe | Resolved | Severe | Resolved | ACZ 4 g | Stopped |
| 13 | 184 | HA, TVO | Completely resolved | Mild | Resolved | Moderate | Resolved | ACZ 250 mg, TPM 25 mg | Stopped |
| 14 | 16.7 | HA, PT | Completely resolved | Severe | Resolved | Severe | Resolved | ACZ 2 g | Stopped |
| 15 | 66.9 | HA, TVO, DP | All decreased | Moderate | Resolved | Moderate | Resolved | TPM 75 mg | Stopped |

ACZ = acetazolamide, DP = diplopia, FU = follow-up after stent placement; HA = headache, Post = post-stent, Pre = pre-stent, PT = pulsatile tinnitus, TPM = topiramate, TVO = transient visual obscuration

Functional and anatomic outcomes are presented in Table 3. VA improved in 9 eyes (30%) of 7 patients, worsened in 4 eyes (13%) of 4 patients, and remained stable in 17 eyes (57%) of 11 patients. Change in VA between the baseline and most recent visit was not significant (0 logMAR, IQR -0.1 – 0, p=0.09). However, VFMD (median increase 3.0, IQR 2.0 – 4.2, p<0.001) and pRNFL swelling (median decrease 147 μm, IQR 41.8 – 242.8, p<0.001) improved significantly after stenting. VFMD improved in 13 eyes (48%, range 3.2 – 16.5 dB) of 9 patients, worsened in 1 eye (4%), and remained stable in 13 eyes (48%) of 10 patients. Among the 8 eyes of 4 patients with fulminant IIH, VA improved in 3 eyes, worsened in 2 eyes, and remained stable in 3 eyes. Change in VA was also not significant (p=0.402). Change in VFMD could be calculated for 6 eyes, of which 3 improved and 3 remained stable.

**Table 3.** Functional and anatomic outcomes before and after stenting.

| Patient | Total FU (weeks) | VA – right eye (logMAR) |  | VA – left eye (logMAR) |  | MD – right eye (dB) |  | MD – left eye (dB) |  | pRNFL – right eye (μm) |  | pRNFL – left eye (μm) |  |
| --- | --- | --- | --- | --- | --- | --- | --- | --- | --- | --- | --- | --- | --- |
|  |  | Pre | Post | Pre | Post | Pre | Pre | Pre | Post | Pre | Post | Pre | Post |
| 1 | 73 | 0.48 | 0.46 | 1.9 | 1.3 | -11.43 | -8.21 | -19.6 | -3.08 | 99 | 69 | 127 | 70 |
| 2 | 49.1 | 0.34 | 0.02 | 0.38 | 0.2 | -4.4 | -0.73 | -4.28 | -2.03 | 113 | 104 | 151 | 114 |
| 3 | 65.9 | 0 | 0.02 | 0 | 0.14 | -19.23 | -13.97 | -12.24 | -9.25 | 322 | 78 | 395 | 89 |
| 4 | 96.4 | 0 | 0 | 0 | 0 | -3.08 | -1.54 | -2.27 | UR | 242 | 96 | 192 | 96 |
| 5 | 55.3 | 0 | 0 | 0 | 0 | -5.98 | -3.29 | -7.29 | -3.13 | 391 | 180 | 184 | 115 |
| 6 | 220.1 | 0.1 | 0 | 1.9 | 2.3 | -21.83 | -19.46 | -33.75 | -33.15 | 58 | 54 | 55 | 45 |
| 7 | 108.4 | 0.18 | 0.04 | 0.1 | 0.06 | -3.88 | -1.9 | -4.35 | -2.29 | 398 | 89 | 324 | 85 |
| 8 | 110.1 | 0.3 | 0.1 | 0.3 | 0.4 | -6.69 | -3.18 | -11.94 | -5.52 | 443 | 64 | 480 | 72 |
| 9 | 174.6 | 0.18 | 0.3 | 0.3 | 0.3 | -14.05 | -6.39 | -11.13 | -6.05 | 331 | 107 | 247 | 99 |
| 10 | 7.4 | 0 | 0 | 0 | 0 | -4.7 | -4.27 | -3.88 | -3.44 | 225 | 181 | 360 | 140 |
| 11 | 23.0 | 1.9 | 0.34 | 0.6 | 0.22 | UR | -24.28 | UR | -25.17 | UR | 93 | UR | 90 |
| 12 | 17.1 | 0 | 0 | 0 | 0 | -7.01 | -3.44 | -6.68 | -4.04 | 383 | 85 | 376 | 94 |
| 13 | 184 | 0.02 | 0 | 0 | 0 | -0.4 | -5.3 | -1.85 | -1.72 | 149 | 108 | 335 | 97 |
| 14 | 16.7 | 0.28 | 0.18 | 0.14 | 0.1 | -2.94 | -0.67 | -6.08 | -1.95 | 107 | 83 | 250 | 83 |
| 15 | 66.9 | 0.18 | 0.18 | 0.18 | 0.18 | -9.57 | -6.04 | -14.65 | -7.95 | 160 | 100 | 187 | 108 |

VA = best-corrected visual acuity, MD = mean deviation on automated perimetry, UR = unreliable

Compared to patients with complete postoperative cessation of symptoms, papilledema, and pharmacotherapy, patients with failed stenting had been affected by IIH (43.6 vs. 26.4 weeks, p=0.69) and associated symptoms (8.3 vs. 3.7 weeks, p=0.86) for a longer duration. However, these differences were not significant. IIH duration and visual symptom duration were not significantly associated with change in VA nor change in VFMD.

The most recent follow-up venogram was performed a median of 20.8 weeks (11.3 – 49.8) after TSS. Postoperative transverse sinus restenosis and/or in-stent thrombosis was not observed in any patient. One patient developed postoperative thrombosis of the vein of Labbe and was treated with rivaroxaban. No other postoperative complications occurred, and no patient required additional surgical intervention for IIH.

## Discussion

### Summary of results

In this case series of patients who underwent TSS for IIH, all patients were able to wean off ICP-lowering pharmacotherapy completely (71%) or to a lower dose (29%). Furthermore, all patients demonstrated improvement or resolution of papilledema and ICP-related symptoms after TSS. While most patients demonstrated stable VA post-TSS, VFMD improved in over 85% of patients. Lastly, there were no major TSS-associated complications, and no patient required a subsequent surgical intervention for IIH. One patient experienced postoperative thrombosis at a remote location from stenting.

### Comparison to previous literature

Our study adds to the current scarcity of evidence on pharmacotherapeutic outcomes after TSS. Previous studies found that 31%-100% of patients can be weaned off ICP-related pharmacotherapy completely after TSS,^13–20^ and that a further 17%-30% can be weaned to lower doses.^13,15,19^ Our study showed similar rates of pharmacotherapy discontinuation (71%) and reduction (29%).

Evidence on visual prognosis after TSS is heterogeneous, which is partially attributable to varying inclusion criteria that studies have imposed on baseline visual function and papilledema status. We conducted a literature review of studies that reported post-TSS visual outcomes, applying our study definition of meaningful VA and VFMD change when individual participant data were reported and the original study’s definition if individual participant data were not available. In studies that enrolled patients with both fulminant and non-fulminant IIH, the rate at which visual acuity improves, remains stable, and worsens after TSS ranges from 8%-87%, 14%-92%, and 0%-21%, respectively.^13,15,18,21–27^ For VFMD, these values are 17%-80%, 9%-83%, and 0%-8%, respectively.^13,15,17,18,21,24,27^ Among patients with fulminant IIH, the rate at which visual acuity improves, remains stable, and worsens after TSS ranges from 25-100%, 0-50%, and 25-100%, respectively.^14,21^ For VFMD, these values are 75-100%, 0%, and 0-25%, demonstrating less heterogeneity than VA outcomes.^14,16,21^ Systematic reviews and meta-analyses have found the rate of VA improvement post-TSS to range from 69% to 78%;^9–11^ one meta-analysis combined VA, VFMD, and subjective visual complaints into a composite outcome and found a postoperative improvement rate of 74%.^12^ However, none of these systematic reviews provided a quantitative definition of VA change. Our study demonstrated a relatively lower rate of VA improvement but higher rate of VFMD improvement, a pattern that was consistent among the fulminant IIH subgroup. Our literature review similarly demonstrated that VFMD tends to show greater and more consistent improvement after TSS than VA, especially among individuals with fulminant IIH. Thus, we propose that VFMD is a better surrogate for TSS efficacy than VA.

Elder et al. analyzed 4 patients with acute vision loss due to IIH.^14^ Inclusion criteria were not clearly described and the patients varied widely with respect to baseline visual function and time to presentation. After TSS, two showed improvement in visual function, one had stable VA but worse VF, and one patient showed deterioration of VA and VF; this patient already had optic atrophy prior to TSS.^14^ Notably, all patients were treated with temporary CSF diversion as a bridge to stenting. Our cohort did not receive temporary CSF diversion and had favorable visual outcomes, even in patients with fulminant IIH. Thus, it is unlikely that temporary CSF diversion procedure is necessary in this subgroup of patient and may adversely increase the risk of spinal cord epidural bleeding since patients undergoing TSS require dual platelet anticoagulation.

All patients in our case series experienced improvement or resolution of ICP-related symptoms and papilledema confirmed by peripapillary OCT. This finding is consistent with previous systematic reviews and meta-analyses which demonstrated high rates of papilledema (86%-97%) and symptomatic improvement (overall 87%, headache 78%-83%, tinnitus 85%-95%) after venous sinus stenting.^8–12^ None of our patients experienced major complications, developed in-stent thrombosis or restenosis, nor required subsequent surgical intervention for IIH. Meta-analyses of patients undergoing venous sinus stenting for IIH found that major and minor complications occur in less than 3% and 5% of cases, respectively, with a mortality rate of 0%.^8–10,12^ Major complications of TSS include intracranial hemorrhage and stroke; minor complications include transient hearing loss, femoral pseudoaneurysm, retroperitoneal hematoma, urinary tract infection, and syncope. The rates of stent survival and stent-adjacent stenosis are estimated to be 84%-100% and 14%, respectively.^9,12^ Subsequent surgical treatment for IIH is required in 10%-13% of patients.^8,10–12^ Compared to ONSF and CSF diversion procedures, venous sinus stenting leads to similar or better rates of visual recovery, symptomatic improvement, complications, and surgical re-treatment.^10,11^ These findings indicate that venous sinus stenting is an efficacious and safe procedure that should be considered in IIH patients who are refractory or intolerant of ICP-lowering pharmacotherapy. Prospective systematic reviews should examine functional, anatomic, and pharmacotherapy outcomes after venous sinus stenting.

### Limitations

We acknowledge some important limitations. First, there are inherent biases to the retrospective nature of this study. Second, our study included a small cohort and is likely subject to selection bias favoring inclusion of patients with more severe disease courses, which is an inevitable by-product of strict indications for TSS. Lastly, our patients had a wide range of baseline visual function.

Our study supports TSS as an effective and safe therapeutic modality for patients with medically refractory or fulminant IIH. VF may be a better indicator of treatment success than VA. Prospective, well-powered studies are needed to ascertain the efficacy and safety of TSS. In particular, a multicentre randomized controlled trial should be conducted to compare TSS against CSF diversion procedures, ONSF, and conservative treatment. The effect of time to treatment on VA and VF parameters should be assessed in future research.

## Data Availability

All data produced in the present study are available upon reasonable request to the authors.

## REFERENCES

1. Friedman DI, Liu GT, Digre KB. Revised diagnostic criteria for the pseudotumor cerebri syndrome in adults and children. Neurology. 2013;81(13):1159–1165.

2. Farb RI, Vanek I, Scott JN, et al. Idiopathic intracranial hypertension: the prevalence and morphology of sinovenous stenosis. Neurology. 2003;60(9):1418–1424.

3. Johnson LN, Krohel GB, Madsen RW, March GA Jr. The role of weight loss and acetazolamide in the treatment of idiopathic intracranial hypertension (pseudotumor cerebri). Ophthalmology. 1998;105(12):2313–2317.

4. NORDIC Idiopathic Intracranial Hypertension Study Group Writing Committee, Wall M, McDermott MP, et al. Effect of acetazolamide on visual function in patients with idiopathic intracranial hypertension and mild visual loss: the idiopathic intracranial hypertension treatment trial. JAMA. 2014;311(16):1641–1651.

5. Corbett JJ, Savino PJ, Thompson HS, et al. Visual loss in pseudotumor cerebri. Follow-up of 57 patients from five to 41 years and a profile of 14 patients with permanent severe visual loss. Arch Neurol. 1982;39(8):461–474.

6. Thambisetty M, Lavin PJ, Newman NJ, Biousse V. Fulminant idiopathic intracranial hypertension. Neurology. 2007;68(3):229–232.

7. Wall M, George D. Idiopathic intracranial hypertension. A prospective study of 50 patients. Brain. 1991;114 (Pt 1A):155–180.

8. Nicholson P, Brinjikji W, Radovanovic I, et al. Venous sinus stenting for idiopathic intracranial hypertension: a systematic review and meta-analysis. J Neurointerv Surg. 2019;11(4):380–385.

9. Saber H, Lewis W, Sadeghi M, Rajah G, Narayanan S. Stent Survival and Stent-Adjacent Stenosis Rates following Venous Sinus Stenting for Idiopathic Intracranial Hypertension: A Systematic Review and Meta-Analysis. Interv Neurol. 2018;7(6):490–500.

10. Satti SR, Leishangthem L, Chaudry MI. Meta-Analysis of CSF Diversion Procedures and Dural Venous Sinus Stenting in the Setting of Medically Refractory Idiopathic Intracranial Hypertension. AJNR Am J Neuroradiol. 2015;36(10):1899–1904.

11. Scherman DB, Dmytriw AA, Nguyen GT, et al. Shunting, optic nerve sheath fenestration and dural venous stenting for medically refractory idiopathic intracranial hypertension: systematic review and meta-analysis. Ann Eye Sci. 2018;3:26–26.

12. Teleb MS, Cziep ME, Lazzaro MA, et al. Idiopathic Intracranial Hypertension. A Systematic Analysis of Transverse Sinus Stenting. Interv Neurol. 2013;2(3):132–143.

13. Dinkin MJ, Patsalides A. Venous Sinus Stenting in Idiopathic Intracranial Hypertension: Results of a Prospective Trial. J Neuroophthalmol. 2017;37(2):113–121.

14. Elder BD, Goodwin CR, Kosztowski TA, et al. Venous sinus stenting is a valuable treatment for fulminant idiopathic intracranial hypertension. J Clin Neurosci. 2015;22(4):685–689.

15. Hendrix P, Whiting CJ, Griessenauer CJ, Bohan C, Schirmer CM, Goren O. Neuro-ophthalmological evaluation including optical coherence tomography surrounding venous sinus stenting in idiopathic intracranial hypertension with papilledema: a case series. Neurosurg Rev. 2022;45(3):2239–2247.

16. Horev A, Ben-Arie G, Walter E, et al. Emergent cerebral venous stenting: A valid treatment option for fulminant idiopathic intracranial hypertension. J Neurol Sci. 2023;452:120761.

17. Kulhari A, He M, Fourcand F, et al. Safety and Clinical Outcomes after Transverse Venous Sinus Stenting for Treatment of Refractory Idiopathic Intracranial Hypertension: Single Center Experience. J Vasc Interv Neurol. 2020;11(1):6–12.

18. Oyemade KA, Xu TT, Brinjikji W, et al. Improved Ophthalmic Outcomes Following Venous Sinus Stenting in Idiopathic Intracranial Hypertension. Frontiers in Ophthalmology. 2022;2. doi:10.3389/fopht.2022.910524

19. Patsalides A, Oliveira C, Wilcox J, et al. Venous sinus stenting lowers the intracranial pressure in patients with idiopathic intracranial hypertension. J Neurointerv Surg. 2019;11(2):175–178.

20. Reid K, Winters HS, Ang T, Parker GD, Halmagyi GM. Transverse Sinus Stenting Reverses Medically Refractory Idiopathic Intracranial Hypertension. Frontiers in Ophthalmology. 2022;2. doi:10.3389/fopht.2022.885583

21. Ahmed RM, Wilkinson M, Parker GD, et al. Transverse sinus stenting for idiopathic intracranial hypertension: a review of 52 patients and of model predictions. AJNR Am J Neuroradiol. 2011;32(8):1408–1414.

22. Bussière M, Falero R, Nicolle D, Proulx A, Patel V, Pelz D. Unilateral transverse sinus stenting of patients with idiopathic intracranial hypertension. AJNR Am J Neuroradiol. 2010;31(4):645–650.

23. Donnet A, Metellus P, Levrier O, et al. Endovascular treatment of idiopathic intracranial hypertension: clinical and radiologic outcome of 10 consecutive patients. Neurology. 2008;70(8):641–647.

24. Smith KA, Peterson JC, Arnold PM, Camarata PJ, Whittaker TJ, Abraham MG. A case series of dural venous sinus stenting in idiopathic intracranial hypertension: association of outcomes with optical coherence tomography. Int J Neurosci. 2017;127(2):145–153.

25. Fields JD, Javedani PP, Falardeau J, et al. Dural venous sinus angioplasty and stenting for the treatment of idiopathic intracranial hypertension. J Neurointerv Surg. 2013;5(1):62–68.

26. Liu X, Di H, Wang J, et al. Endovascular stenting for idiopathic intracranial hypertension with venous sinus stenosis. Brain Behav. 2019;9(5):e01279.

27. Radvany MG, Solomon D, Nijjar S, et al. Visual and neurological outcomes following endovascular stenting for pseudotumor cerebri associated with transverse sinus stenosis. J Neuroophthalmol. 2013;33(2):117–122.

